# Identifying Opportunities for Fluid Balance Optimization in Critically Ill Children

**DOI:** 10.1101/2025.01.12.25320399

**Authors:** Denise C. Hasson, Ami Shah, Chloe G. Braun, Ulka Kothari, Steve Drury, Heda Dapul, Julie C. Fitzgerald, Celeste Dixon, Andrew Barbera, James Odum, Nina Terry, Scott L. Weiss, Susan D. Martin, Adam C. Dziorny

## Abstract

**Introduction:** Fluid overload (FO), a state of pathologic positive cumulative fluid balance (CFB), is common in Pediatric Intensive Care Units (PICU) and associated with morbidity and mortality. Because different PICUs may have unique needs, barriers, and limitations to accurately report fluid balance (FB) and reduce FO, understanding the drivers of positive FB is needed. We hypothesize CFB >5% and >10% is common on ICU days 1 and 2, but that reasons for high %CFB will vary across sites, as will barriers to accurate FB recording and opportunities to improve FB recording/management.

**Methods:** Concurrent mixed methods study utilizing a retrospective observational cohort design and prospective interview and survey design performed at four tertiary pediatric ICUs. FB data was extracted from the electronic health record. A federated data collection framework allowed for rapid data aggregation. The primary outcome was %CFB on ICU days 1 and 2, defined as total intake minus total output divided by ICU admission weight. Chi-square test and Wilcoxon rank sum tests compared results across and within sites.

**Results:** Amongst 3,071 ICU encounters, day 2 CFB >5% varied from 39% to 54% (p=0.03) and day 2 CFB >10% varied from 16% to 25% (p=0.04) across sites. Urine occurrence recordings and patients receiving >100% Holliday-Segar fluids on Day 1 differed across sites (p<0.001). Sites discussed overall FB and specific FB goals on rounds with differing frequency (42-73% and 19-39%, respectively), but they reported similar barriers to accurate FB reporting and achievable opportunities to improve FB measurements, including patients/families not saving urine/stool, patients not tracking oral intake, and lack of standardized charting of flushes.

**Conclusion:** Day 2 CFB >5% and >10% was common among pediatric ICU encounters but proportion of patients varied significantly across ICUs. Individual ICUs have different drivers of FO that must be targeted to improve FB management.

## Introduction

Fluid overload (FO), a state of pathologic positive cumulative fluid balance (CFB), is common in children with critical illness and is associated with prolonged mechanical ventilation, increased risk of acute kidney injury (AKI), increased intensive care unit (ICU) length of stay (LOS), morbidity, and mortality.^1^ Although early literature associating FO with morbidity and mortality focused on critically ill children requiring renal replacement therapy (RRT),^2–5^ most pediatric ICU (PICU) patients with FO have similar associations with poor outcomes.^6^ A 2018 meta-analysis found that %CFB >5% within 24 hours of PICU admission was associated with nine-fold greater odds of mortality, while peak %CFB >10% during PICU admission was associated with 15-fold greater mortality.^1^ Awareness of this excess mortality has led to awareness of the need for unified definitions^1^ as well as increased interest in strategies to prevent, mitigate, and reduce positive %CFB.

Recent international multicenter studies that have investigated the impact of amount and temporality of this positive %CFB showed that small amounts (≥5%) early in the ICU course (Day 1 and Day 2) are associated with the aforementioned negative ICU outcomes.^1,6^ One approach to solving this problem is to develop a clearer understanding of the key drivers of fluid exposure.^6^ Studies have begun to identify reasons behind positive %CFB, such as fluid creep (i.e., the administration of non-nutritive, non-resuscitative fluids)^7^ and have found associations between this metric and negative PICU outcomes.^8,9^ However, we lack a holistic, multidisciplinary, and multicenter approach to investigating this problem, including identification of other key drivers of fluid exposure, barriers to accurate fluid balance (FB) reporting, and targeted fluid deresuscitation.

Nuances in FB management may vary across institutions and even between units within the same institution due to practice variability and operational differences that come from size and geographic diversity. Unique challenges to optimizing fluid sensitive care include accurate FB reporting, lack of structured discussion and goal setting around FB, and varied opportunities and actions to reduce %CFB. The objective of this study was to quantify the frequency of positive %CFB amongst critically ill pediatric patients in our multicenter collaborative and qualitatively identify areas for improvement of fluid sensitive care. We hypothesize that %CFB >5% and >10% is common within initial days of pediatric ICU admission, but that reasons for high %CFB will vary across sites, as will barriers to accurate FB recording and opportunities to improve FB recording and management.

## Materials and Methods

### Study Design

This is a concurrent mixed methods study conducted within a collaborative composed of four tertiary care centers (including four PICUs and two pediatric cardiac ICUs (CICU)) spanning 185 pediatric ICU beds. We performed a quantitative retrospective observational cohort study to measure FB metrics and ICU outcomes from all new pediatric ICU admissions in the 6 months prior to qualitative data collection, between April 2022 and January 2023. All sites use the same Electronic Health Record (EHR), Epic (Epic Systems Corporation, Verona, WI). Site A included encounters from their 12 bed PICU and 15 bed CICU. Site B has included encounters from their 22 bed PICU. Site C has included encounters from their 74 bed PICU and 38 bed CICU. Site D has included encounters from their 24 bed PICU. All PICUs are mixed medical/surgical units, and all can provide extracorporeal support (renal replacement therapy, extracorporeal membrane oxygenation). Of note, Site B is not a trauma center.

We also performed prospective observational surveys using qualitative methods, including interviews and surveys with bedside nurses to understand current ICU-specific practices and barriers. This study followed procedures in accordance with ethical standards as delineated by the Helsinki Declaration of 1975. Each site’s institutional review board deemed the project exempt from IRB review.

We used a federated data collection framework to collect and aggregate clinical data.^10^ We wrote data extraction queries, analysis scripts, survey data dictionaries, and semi-structured interview scripts at a single site and disseminated each to all sites to promote process standardization. Each site’s data remained at its original site; however, because the format was standardized, the same analysis scripts were run at each site simultaneously. Aggregated outputs were shared among sites and statistical comparisons were computed from these aggregate outputs. This allowed for site-specific understanding of FB management while still allowing for comparison and collaboration. No row-level (i.e., individual patient level) data were shared between sites.

### Baseline Measures and Definitions

We collected metrics including baseline demographics, admission weight, height, total intake and output by route and type, urine occurrence counts (e.g. urine, stool, or mixed output that was not weighed or measured as an exact volume), number of patients with invasive mechanical ventilation orders (as a surrogate for use of invasive mechanical ventilation), and ICU LOS and hospital disposition (including mortality). Total intake and output included all measures noted to be “intake” or “output” within the EHR. We manually harmonized input and output subtypes among sites where necessary in order to identify intravascular fluid subtypes and urine output subtypes, to ensure subsequent calculations were consistent across sites.

Consistent with prior studies, we attributed fluid measures to ICU days as follows: Day 0 was defined as admission through the next 7 AM, and each subsequent ICU day (1 through 7) included all fluid measures from 7 AM through the next 7 AM or ICU discharge.^6,11^

We calculated %CFB as:^12^

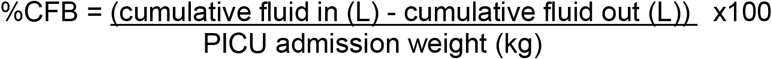

We calculated the ratio of fluids administered to Holliday-Segar (H-S) maintenance fluid rates as: (the sum of all IV fluids including parenteral nutrition) / (4x first 10 kg weight + 2x number of kg between 10-20 + 1x number of kg >20).^13^ Of note, each site uses the same EHR, and all Sites had admission orders that contained maintenance fluid rates that were calculated as 100% of H-S fluid volume, although Site C provided options for 100%, 66%, and 150%. Ultimately the maintenance fluid rate was modifiable and decided upon by the medical team. All IV fluids included any intake not labeled as enteral, blood product, extracorporeal (e.g., CRRT), or “other” and included both prescribed fluids, medications, and IV flushes.

### Interview and Survey Based Assessment of ICU FB Practice

We followed a multistep survey creation framework to identify site-specific barriers and facilitators to improve accurate FB reporting.^14^ We wrote a question script based on goals we hoped to achieve (**Supplementary Methods**), then we interviewed 3-5 nurses at Sites A and B. These answers informed the questions and responses that we incorporated into the Research Electronic Data Capture (REDCap) Nurse Survey (**Supplementary Methods**). After drafting specific questions with informed multiple-choice answers, we emailed surveys to bedside nurses at each site and allowed them a month to complete this survey. We shared data dictionaries and implemented the same survey in each site-specific REDCap instance. Nurses were asked to choose the three best answers for questions 1 and 2, and the single best answer for question 3 of the REDCap survey.

We measured the frequency and content of discussions related to FB, recognizing that providers cannot address an issue they 1) are unaware of and 2) do not discuss. We focused on discussions during ICU rounds, as this is when daily plans and goals are established in collaboration with a multidisciplinary team. Study team members (Sites A, B) or research assistants trained by study team members (Site C) interviewed available nurses 1-4 hours after rounds (**Supplementary Methods**) 2-3 times weekly for 4-6 weeks (to account for the varied practices of different providers). Site D employed a quality improvement-focused research assistant that observed daily PICU rounds (Monday-Friday) to record the frequency of FB discussions for 6 weeks. We recorded observation results in a standard REDCap instance or shared Excel spreadsheet (Microsoft, Redmond, WA, USA) and aggregated results across sites.

### Statistical Analysis

We performed clinical data analyses at each individual site using analysis scripts written in R (https://www.r-project.org, R Foundation for Statistical Computing, Vienna, Austria). Results are presented by institution. We reported categorical data in frequencies (%) over a given period (e.g., one week or 7 days) and aggregate data as median (interquartile range, IQR). We compared counts across and within sites using Chi-square and compared distributions within sites using Wilcoxon rank sum tests. Baseline fluid metric comparisons by site excluded patients with urine “occurrences,” as such occurrences suggest that urine or stool volume was not recorded, thus biasing toward a more positive %CFB than what may have occurred. However, we also performed sensitivity analyses including encounters with urine occurrences without calculating or extrapolating missing data. We reported descriptive statistics of survey results at each site and aggregated them across all sites.

## Results

### Site Specific Demographics and Outcomes

Over 6 months, we included 3,071 ICU encounters and 2,850 hospitalizations encompassing 2,632 unique patients (**Table 1** and **Supplementary Table S1**). Median age at ICU admission ranged from 3.6 to 7.4 years old, and 41-49% of patients at each site were female. Race and ethnicity varied by site but the predominant race was White/Caucasian at each site, and Hispanic or Latino ethnicity ranged from 7-22% of each cohort. ICU LOS was between 35-55 hours, and mortality rates were between 0.7-3.5%, although this difference was due to varied disposition to other facilities (**Supplementary Table S1**).

**Table 1:** Baseline Fluid Metric Comparisons by Site, Not Including Urine Occurrences

| Fluid Metric | Overall<br>(N=3071) | Site A<br>(N=427) | Site B<br>(N=553) | Site C<br>(N=1356) | Site D<br>(N=735) | Chi Square<br>Test |
| --- | --- | --- | --- | --- | --- | --- |
| Day 1 CFB<br>>5%** | 697/2064<br>(34%) | 61 / 178<br>(34%) | 133 / 427<br>(31%) | 344 / 950<br>(36%) | 159 / 509<br>(31%) | X<sup>2</sup>=5.32<br>P=0.15 |
| Day 1 CFB<br>>10%** | 205/2064<br>(9.9%) | 14 / 178<br>(7.9%) | 45 / 427<br>(11%) | 96 / 950<br>(10%) | 50 / 509<br>(9.8%) | X<sup>2</sup>=1.06<br>P=0.79 |
| Day 2 CFB<br>>5%** | 670/1437<br>(47%) | 48 / 89<br>**(54%)** | 113 / 287<br>**(39%)** | 351 / 732<br>(48%) | 158 / 329<br>(48%) | **X<sup>2</sup>=8.75**<br>**P=0.03** |
| Day 2 CFB<br>>10%** | 292/1437<br>(20%) | 22 / 89<br>(25%) | 46 / 287<br>**(16%)** | 166 / 732<br>(23%) | 58 / 329<br>**(18%)** | **X<sup>2</sup>=8.31**<br>**P=0.04** |
| Urine<br>Occurrences | 2017/9324<br>(22%) | 667 / 1315<br>**(51%)** | 292 / 1618<br>(18%) | 694 / 4479<br>(15%) | 364 / 1912<br>(19%) | **X<sup>2</sup>=775.8**<br>**P < 0.001** |
| Day 1 H-S<br>fluids >100% | 712/2510<br>(28%) | 66 / 335<br>**(20%)** | 124 / 433<br>(29%) | 341 / 1096<br>(31%) | 181 / 646<br>(28%) | **X<sup>2</sup>=16.5**<br>**P < 0.001** |
| Day 2 H-S<br>fluids >100% | 366/1461<br>(25%) | 40 / 190<br>(21%) | 65 / 235<br>(28%) | 188 / 709<br>(27%) | 73 / 327<br>(22%) | X<sup>2</sup>=4.58<br>P=0.21 |

Cumulative fluid balance (CFB), urine occurrence measurements, and IVF intake above H-S calculation for each site. Site-specific sample size in header row reflects total ICU admissions over the study period. Each cell reports number of patient-days (%) meeting fluid metric. Cell-specific denominator reflects total number of ICU patients present on that day of ICU stay, or during any day if no day is specified. Differences between cell specific denominators and total site denominator reflect patients excluded due to having urine occurrences. H-S fluids metric only includes patients with IVF intake. We report row-wise Chi-square p-value to examine the relationship between fluid metric and site across all four sites. \*\* indicates only those samples without urine occurrences. When the chi-square test resulted with a significant association, we subjectively labeled the outliers blue to indicate a favorable difference and red to indicate an unfavorable difference. P-values <0.05 represented with bolding. CFB-cumulative fluid balance, H-S- Holliday-Segar.

### Fluid Balance Metric Comparisons by Site

Excluding patients with urine occurrences charted, CFB >5% was common early in the ICU course, occurring in 34% (N=697/2,064) of ICU encounters on Day 1 and 47% (N=670/1,437) of ICU encounters on Day 2 (**Table 1**). Ten percent (N=205/2,064) of ICU encounters had >10% CFB on Day 1 and 20% (N=292/1,437) had >10% CFB on Day 2. Total number (%) of ICU encounters with Day 2 CFB >5% and >10% significantly differed by site (**Table 1**, p=0.03 and p=0.04, respectively). Day 1 CFB >5% and >10% did not differ between sites (**Table 1**, p=0.15 and p=0.79, respectively), with sites having 5% or less difference between incidence of CFB >5% and 3% or less difference for CFB >10%. However, when patients with urine occurrences were included, day 1 CFB >5% also differed by site (**Supplementary Table S2**).

Instead of measuring all total IVF volume measured (excluding enteral nutrition and blood but including parenteral nutrition, as some EHRs do not differentiate the latter from IVF), we chose to measure total number (%) of ICU encounters where patients’ total IVF was >100% of H-S fluid volume. Sites varied in the number of ICU encounters where >100% of H-S fluids were prescribed on Day 1 but not on Day 2 (**Table 1**, p < 0.001 and p=0.21, respectively). All sites had at least 20% of encounters where >100% H-S fluids were prescribed on both days, but on day 1, multiple sites approached 30% of encounters with >100% H-S fluids prescribed.

Given the importance of accurate FB recording to these metrics, we measured the number of encounters that had a urine occurrence documented. The frequency of encounters with urine occurrences varied by site and ranged from 15.5% to 50.7% (**Table 1**, p <0.001).

Mechanical ventilation orders were more common in those encounters that had >100% of H-S fluids prescribed at all sites on Day 1 (A: 28.5% vs 13.3%, p<0.001; B: 40.3% vs 23.7%, p=0.001; C: 49.8% vs 23.4%, p<0.001; D: 43.9% vs 21.0%, p<0.001). Mechanical ventilation orders were more common in those with >100% of H-S fluids prescribed at sites C and D on Day 2 but not sites A and B (A: 25.0% vs 15.4%, p=0.11; B: 34.0% vs 23.4%, p=0.07; C: 43.0% vs 15.8%, p<0.001; D: 33.8% vs 12.5%, p<0.001) at each site (**Table 2**). The lack of difference seen for sites A and B on Day 2 could have been partially due to smaller sample sizes on this day; however, it is worth noting that Site A had a lower number of encounters with >100% of H-S fluids prescribed in both groups compared to the other sites.

**Table 2:** Frequency of Encounters with >100% Holliday-Segar Fluids by Mechanical Ventilation Status, by Site

|  | Site A |  | Site B |  | Site C |  | Site D |  |
| --- | --- | --- | --- | --- | --- | --- | --- | --- |
|  | +MV | -MV | +MV | -MV | +MV | -MV | +MV | -MV |
| ICU Day 1, N (%) | **40/140**<br>**(28.6%)** | **26/195**<br>**(13.3%)** | **52/129**<br>**(40.3%)** | **72/304**<br>**(23.7%)** | **159/319**<br>**(49.8%)** | **182/777**<br>**(23.4%)** | **87/198**<br>**(43.9%)** | **94/448**<br>**(21.0%)** |
| ICU Day 2, N (%) | 28/112<br>(25.0%) | 12/78<br>(15.4%) | 32/94<br>(34.0%) | 33/141<br>(23.4%) | **120/279**<br>**(43.0%)** | **68/430**<br>**(15.8%)** | **51/151**<br>**(33.8%)** | **22/176**<br>**(12.5%)** |

We report the n (%) of patient encounters with >100% H-S fluids in patients who had a mechanical ventilation order (+MV) and the n (%) of patient encounters with >100% H-S fluids in patients who did not have a mechanical ventilation order (-MV). Chi Square test was used to compare the association of >100% H-S fluids based on MV status on each ICU day, by site. Bolded values indicate comparisons are statistically significant with a p-value < 0.05, though all significant p-values were < 0.001. MV-mechanical ventilation, ICU-intensive care unit.

In unadjusted analyses excluding patients with urine occurrences, ICU LOS was significantly longer in patients with Days 1 and 2 CFB >5% at all sites (**Table 3**, p < 0.01). ICU LOS was significantly longer in patients with Days 1 and 2 CFB >10% at three of four sites, associated with a median of 1.5-2.5 greater ICU days (**Table 3)**.

**Table 3:** Intensive Care Unit Length of Stay by % Cumulative Fluid Balance Metric, by Site

|  | Site A |  | Site B |  | Site C |  | Site D |  |
| --- | --- | --- | --- | --- | --- | --- | --- | --- |
|  | > CFB | < CFB | > CFB | < CFB | > CFB | < CFB | > CFB | < CFB |
| ICU Day 1, 5 % cut-off | **4.5**<br>**(2.2,8.6)** | **1.0**<br>**(0.6,3.6)** | **2.2**<br>**(1.6,4.2)** | **1.6**<br>**(0.9,2.8)** | **2.9**<br>**(1.7,6.2)** | **2.1**<br>**(1.1,4.8)** | **2.6**<br>**(1.4,5.9)** | **1.3**<br>**(0.8,2.6)** |
| ICU Day 1, 10% cut-off | 3.2<br>(2.3,5.8) | 1.9<br>(0.8,4.9) | 3.5<br>(2.0,9.2) | 1.8<br>(1.0,2.8) | **4.1**<br>**(2.4,6.9)** | **2.3**<br>**(1.1,4.9)** | **3.1**<br>**(1.9,7.1)** | **1.5**<br>**(0.9,3.0)** |
| ICU Day 2, 5 % cut-off | 4.7<br>(2.5,8.3) | 3.7<br>(1.6,4.9) | **2.8**<br>**(1.8,5.2)** | **1.9**<br>**(1.3,3.3)** | **3.8**<br>**(2.1,7.4)** | **2.5**<br>**(1.6,5.1)** | **3.3**<br>**(1.9,7.1)** | **2.0**<br>**(1.3,3.6)** |
| ICU Day 2, 10% cut-off | 4.7<br>(2.5,6.9) | 4.0<br>(1.8,6.3) | 4.0<br>(2.2,9.2) | 1.9<br>(1.5,3.4) | **4.3**<br>**(2.8,7.7)** | **2.7**<br>**(1.6,5.4)** | **4.9**<br>**(2.7,8.9)** | **2.2**<br>**(1.4,4.1)** |

Median (interquartile range) length of stay in days for patients who did not meet cut-offs of 5% and 10% CFB compared to those who did, by site, excluding urine occurrences. Wilcoxon rank sum tests determined P values, with P values <0.01 represented by bolding, P value <0.05 represented with *italics*. CFB-cumulative fluid balance, LOS-length of stay.

### Barriers and Opportunities to Improve Accurate FB Recording

All sites had the same top two Most Frequent Barriers to Accurate FB Recording (**Figure 1a, Supplementary Table S3**): “ambulatory patients not saving urine/stool,” and “patients not keeping track of oral intake.” Site D respondents reported that “standardizing charting of medication flushes could be helpful,” while the other three sites reported “families throwing diapers away before being weighed” as an important barrier. All four sites had two of the same four Achievable Opportunities to Accurate FB Recording (**Figure 1b, Supplementary Table S3**): “standardizing recording of flush volumes” and “collaborating with other units (like the emergency department and operating rooms) to add fluids into flowsheets.” Three of four sites shared the other top two opportunities: “standardizing flush volumes per medication” and “consistent zero-ing of bed scales for daily weights.” When asked the most Feasible Method for FB Recording, all sites agreed that “setting a fluid balance goal on rounds and discussing ways to meet it” would be more feasible than “recording strict I/O without urine occurrence measurements” or “obtaining daily weights” (**Figure 1c, Supplementary Table S3**).

### Setting FB Goals

Sites surveyed nurses on a median of 8.5 (8.0-46.5) days (total of 84 days) over a median of 6 (4.25-9.25) weeks (total of 26 weeks) (**Table 4**). All sites discussed FB on 62% (N=578) of patients’ rounds, with Site C discussing FB most frequently (78%, N=93), p<0.001. A FB goal was only set on 23% (N=214) of patient rounds recorded, with Site B setting a goal most frequently (39%, N=26), p=0.005. Sites A and C discussed a FB goal just over 20% of the time (23%, N=52 and 21%, N=103). Sites A-C recorded whether changes were made to the FB plan, such as decreasing maintenance fluids or starting/changing diuretics. Changes were only made for 18% of patients (N=79/439), with sites making changes on 14-36% of patients with varying frequency (p<0.001).

**Table 4:** Assessment of Fluid Balance Practices on Rounds

|  | Site A<br>(N=252) | Site B<br>(N=67) | Site C<br>(N=120) | Site D<br>(N= 494) | Total<br>(N=933) | P<br>value |
| --- | --- | --- | --- | --- | --- | --- |
| Number of Days/Weeks<br>Surveyed | 9 d/ 7 wk | 8 d/ 5 wk | 8 d/ 4<br>wk | 59 d/ 10<br>wk | 84 d/ 26<br>wk |  |
| Discussed Fluid Balance, N (%) | 102 (40) | 46 (69) | 93 (78) | 337 (68) | 578 (62) | <0.001 |
| Set a Fluid Balance Goal, N (%) | 52 (21) | 26 (39) | 33 (28) | 103 (21) | 214 (23) | 0.005 |
| Made Fluid Balance Changes, N (%) | 35 (14) | 24 (36) | 20 (17) | n/a | 79 (18) | <0.001 |

Results of rounds' observations or post-rounds interviews of nurses to report discussions about fluid balance practices and goals. The N reported signifies the number of beds queried or for which rounds were observed, per Site. Site A included data from both pediatric and cardiac intensive care units. Site D did not gather data on whether there were changes made to the plan to address fluid balance goals. d-days, wk-week(s). We report row-wise association of fluid balance discussion and site using Chi-square test.

## Discussion

Across four tertiary pediatric ICUs, CFB >5% and >10% on PICU Days 1 and 2 were very common, with similar rates on Day 1 but varied rates on Day 2 across sites. Sites prescribed total IVF in excess of H-S calculations in 20-30% of ICU encounters, with varying frequency on Day 1 but not on Day 2. Sites routinely discussed FB on rounds, but setting goals and making changes based on said goals occurred <40% of the time. A major limitation to the study of FB is accuracy of FB measuring and recording. We identified that up to 50% of encounters can have urine occurrences charted, leading to inaccurate FB calculations. More importantly, though, we identified many barriers to accurate FB recording and opportunities to improve FB recording, with commonality between sites, but with enough variation to warrant site-specific targeted management.

We have sought to go beyond FB epidemiology to investigate reasons for positive %CFB and to identify barriers to accurate FB measurement and recording so that next steps can be to improve FB recognition. However, we needed to start by understanding our cohorts and their distinct practices. Despite differences in location and size, our centers had similar rates of positive %CFB, which matched the rates of positive %CFB reported by others. In a multicenter international collaborative study, Selewski et al identified CFB >5% on Day 1 and 2 in 38.1% and 53.3% of patients and CFB >10% on Day 1 and 2 in 11.7% and 25.1% of patients.^6^ Another two-center study found Day 3 CFB >10% in 24.6% of patients,^8^ similar to Day 2 %CFB at 2 of our sites. Both of these studies identified strong associations between these FB cut-offs and fluid in excess of hydration requirements with fewer ventilator-free days, ICU-free days, and 28-day ICU mortality.^6,8^ Although these aggregate data greatly add to the known epidemiology of positive %CFB, especially as they pertain to outcomes, they do not discuss intra-unit variability. Specifically, they suggest that next steps should be to identify key drivers of such high %CFB, as this can inform steps to reduce this incidence.^6^

Targeting “fluid creep,” the administration of non-nutritive, non-resuscitation fluid in excess of maintenance fluid requirements, is a worthy strategy for FO prevention given its association with increased morbidity and mortality.^8,9^ Barhight et al found that each 10mL/kg of fluid given in excess of hydration increases the odds of death by 1%.^8^ A single center PICU was able to increase ventilator- and PICU-free days by restricting maintenance fluids to 40% compared to 70-80% in mechanically ventilated children.^15^ Although we have not specifically targeted fluid creep in this study, our data suggest that some sites have more “fluid creep” than others, but that still nearly a quarter of patients may be receiving more fluid than is routinely recommended. This number is lower than others have reported, especially given certain populations are at higher risk for FO than others,^8,9^ but we believe it is still too high given its association with adverse outcomes. Our team of bedside nurses have identified that setting a total fluid goal on rounds and discussing ways to meet it, would be the most feasible method for accurate FB recording. Additionally, our data suggest that more work must be done to improve goal setting, and providers must be cognizant of making adjustments to meet those goals.

It is possible that a major barrier to improving FO incidence may be the difficulty in clinically recognizing positive %CFB. Even 15% CFB can be missed routinely by PICU providers.^7^ Providers must then rely on more objective recorded data to know a patient’s %CFB. Although advances in the EHR have brought extreme amounts of data to the clinicians’ fingertips, it is dependent on and limited by the accuracy of the input data. Simple calculations can be run by the EHR to alert clinicians to unseen FO, but these are made inaccurate by modifiable barriers such as urine occurrences and unclear processes for charting fluids such as medication flushes, as well as non-modifiable barriers such as insensible fluid losses.^16^ Although using daily weights can account for insensible losses,^4^ there are barriers to accurate daily weights in PICU patients, and our data suggest this may be difficult to incorporate into unit workflow. Technical barriers exist to building custom EHR-based scripts that perform complex but clinically useful calculations, such as %CFB. Overcoming these barriers may promote earlier clinical recognition of FO.

There are many strengths to this study. We established a robust multicenter collaborative to aggregate the large amount of granular data necessary for the study of FB without time-intensive manual chart review. Our FB metrics consider inaccuracies that occur when urine and stool are collected as occurrence counts (rather than measured volumes). By comparing practices between sites, we provide feasible solutions that address the nuances of FB management across sites. Our sites have size and geographic diversity, enhancing generalizability. We have leveraged the expertise of our bedside nurses, who often know the most about how much fluid goes into and out of patients and from which sources.

There are also study limitations. Our baseline data was retrospective, and we did not collect data on severity of illness to adjust for in our analyses; thus, differences between sites could have been due to varied patient populations. It is possible that patients with urine occurrences may reflect patients who are less critically ill and, therefore, in whom these inaccuracies are less clinically relevant. To account for this, we performed our retrospective analyses both including and excluding these encounters. To minimize the number of site-specific modifications and parameters within our shared queries and analysis platform, we were unable to collect granular respiratory support data and instead collected ventilator orders as a surrogate for mechanical ventilation. It is for similar reasons around site-specific modifications that we were unable to measure severity of illness markers. We also did not share row-level data among sites, which limited our ability to compute multivariable regression models across sites. Several sites included both PICU and CICU patients, including some mixed within each unit, which may reflect different fluid practices and management strategies. We are intrigued by the similar results reported by pure PICU populations;^6^ future studies should compare fluid practices and resultant fluid creep/%CFB between these populations. We were also unable to differentiate resuscitation fluids from maintenance IVFs, which is why we report IVF as >100% H-S rather than “above maintenance,” per se. As with any survey, there was likely selection bias in those subjects who chose to respond. We attempted to overcome this limitation by including multiple sites to recruit a varied nursing population, but biases may have been present within each site. For the quantification of FB goal setting based on rounds’ discussions, each Site’s results were highly provider dependent. For this reason, an effort was made to observe multiple days across different weeks, to catch a sampling of provider practices. Site D, who used quality improvement aides to observe rounds, may have different results due to their different methods.

Next steps involve measuring acute kidney injury (AKI) frequency to better understand the interplay between AKI and FB in our population. We plan to implement site-specific opportunities to improve FB recording in order to allow for improved recognition of FO. We believe this is a critical step to targeting patients who will benefit most from CFB reduction strategies. We will also be implementing novel EHR-based calculations across sites to assist all team members in identifying positive %CFB earlier.

## Conclusion

Across our multisite collaborative, CFB >5% and >10% was common within the first 2 PICU days but varied across sites. Workflow practices to address positive %CFB varied significantly across sites despite similar barriers to accurate FB recording and opportunities to reduce %CFB. Such opportunities will be the targets of upcoming prospective interventional trials to reduce %CFB and assess ability to improve ICU outcomes.

## Data Availability

All data produced in the present study are available upon reasonable request to the authors.

## Supplementary Materials

Supplementary Methods (PDF)

Supplementary Tables (PDF)

Supplementary Table S1. Site Specific Demographics and Outcomes

Supplementary Table S2. Baseline Fluid Metric Comparisons by Site, Including Urine Occurrences

Supplementary Table S3. Barriers and Opportunities for Accurate Fluid Balance Recording

## Acknowledgements

We would like to acknowledge the work of Denise Dauterman (NYU), who aided in the creation of the fluid balance score, as well as the ICU nurses who completed our surveys and questionnaires and not only provide excellent patient care, but always share their knowledge and insights that have gone into improving our management of FB. We would also like to acknowledge Sheel Chitre and Adrian La Duca (NYU), Amina Khan (CHOP), and Heather Dutton (UAB) for their assistance with clinical data extraction.

## Author Contributions

DH, SM, and ACD conceived of the study and along with AS, CGB, UK, HD, JCF, CD, AB, JO, and SLW contributed to the overall design. ACD and SD wrote software queries and analysis scripts, which were executed at each site by UK, ACD, SD, and JO. All authors contributed to qualitative data collection. ACD and DH completed the analysis and created the figures and tables. DH drafted the manuscript. All authors provided revisions and approved of the final manuscript version.

## Statements and Declarations

### Ethical considerations

This study followed procedures in accordance with ethical standards as delineated by the Helsinki Declaration of 1975. Each site’s institutional review board either approved this study with a waiver of informed consent or deemed the project exempt from IRB review.

### Consent to participate

Waived as above

### Consent for publication

Not applicable

### Data Availability

Data can be made available upon request to corresponding author.

