## Supplementary Information for "Identifying Opportunities for Fluid Balance Optimization in Critically Ill Children"

Supplementary Methods

*Post-Round Assessment of ICU FB Practice Survey Questions*

Research Team members used this script to ask questions to bedside nurses. Answers to the following questions were recorded by patient (e.g. if a nurse were providing care to two patients, they would answer each question separately for each patient/bed space).

1. Did you talk about cumulative balance today? 0-no 1-yes
2. Did you set a fluid balance goal today? 0-no 1-yes
3. Did you make any changes to your plan based on the goal? 0-no 1-yes
4. Patient Dx/Miscellaneous/Comments

*Questions for Development of Nursing Survey for Fluid Balance Process Improvement*

Nursing Questions:  How to improve the accurate capture and incorporation of fluid balance in critically ill patients.

Proposed Script: Thanks for agreeing to participate in this quick set of questions. We are working on project focusing on fluid balance in critically ill patients. As part of this process, we are developing a short survey for bedside nurses, so we can better understand your practice. To help us with this survey we were hoping to ask you a few questions. We’re going to write down the themes of your answers and, combined with others both here and at other institutions in our collaborative, we will develop that brief survey. Because this is a survey there is no IRB approval, but you should know that you don’t have to answer any question and we can stop this discussion at any point! Thanks again for your interest and participation.

(Goal of the question: do nurses think of fluid balance as a helpful data point in managing critically ill patients/help with buy-in) - How do you find fluid balance in PICU patients to be meaningful contributor to clinical status?

(Goal of question: assess if nurses feel like current capture/tools are accurate): Do you feel we accurately capture fluid balance? Are there a group of patients we do better or worse in?

(Goal of the question: what do nurses need to accurately capture fluid balance) - What are some tools that have been, or would be, helpful in accurately capturing a patient's fluid balance?

(Goal of the question: guide our educational ppt) - Are there any topics about fluid balance and its role in critical illness that you think would be helpful to learn more about?

(Goal of the question: ascertain if and what nurses understand about fluid balance) - When asked about fluid balance by the medical team, what do you think about?

(Goal of the question: understand barriers to collecting I/Os/whose involved): How does capturing I/Os affect your daily work flow?  Are there other people involved?

(Goal of the question: understand barriers to current practices that could use improvement or revision): Are there current parts of your workflow for capturing I/O that would benefit from changes or revisions to the workflow? In other words, what are the challenges and how could we fix or improve them?

*PDSA 2: REDCap Fluid Balance Nurse Survey*

Fluid Balance QI Survey: Nurse and Provider Perspectives We appreciate your participation in this short, anonymous, 4-question survey to help guide our quality improvement project. If you have questions, comments, or concerns, please reach out to [team members] directly.  Thank you!

What is your role in the ICU?

Nurse

Provider

Level of Experience in the PICU

< 1 year

1-5 years

5-10 years

10+ years

Which shifts do you work?

Day Shifts

Night Shifts

Mixed/Both

Travel/Per Diem

Which degrees/certifications do you hold?

RN

NP

CNL

CNS

CCRN

Bachelor's

Master's

What are the most frequently encountered barriers to accurately capturing a patient's fluid balance? (Select your top 3 choices)

Too many other tasks de-prioritize fluid balance capture

Patient acuity precludes strict I/O tracking

Perception that it is not important to the clinical team and not worth the time investment

Ambulatory patients do not remember to save urine or stool output

Patients allowed to PO do not keep track of oral intake

Diapers were thrown away by others before I could weigh them

Unclear process for charting medication flushes of varying amounts

Enteral intake is less important to track

Technical challenges with pump integration

Other

What other barriers exist to accurate fluid balance documentation? __________________________________

What are some achievable opportunities to improve accurately capturing a patient's fluid balance? (Select your top 3 choices)

Standardized flush volumes per medication

Standardized recording of flush volumes

Maintaining foley catheters longer for strict I/O

Alternatives to foley catheters such as pure wick or condom catheters

Consistent zeroing of beds for daily weights

Collaboration with non-ICU teams (e.g., emergency department, floor, transport, operating room) to incorporate fluids into flowsheets

Larger scales to allow for accurate weight of adult diapers and bed sheets

Other

What other achievable opportunities exist to accurately capture a patient's fluid balance? __________________________________

If we were to agree that accurate daily cumulative fluid balances are necessary, which of these methods do you feel is most easily achievable? (Select only one choice)

Daily weights in high-risk patients

Strict I/O without occurrence measurements (e.g., stool x1, urine/stool x6)

Setting a total fluid goal daily on rounds and discussing ways to meet it

Supplementary Tables

**Supplementary Table S1**. Site Specific Demographics and Outcomes.

|  | **Site A (N=427)** | **Site B (N=553)** | **Site C (N=1356)** | **Site D**  **(N=735)** |
| --- | --- | --- | --- | --- |
| Unique patients, n | 345 | 450 | 1189 | 648 |
| Unique hospital encounters, n | 388 | 509 | 1263 | 690 |
| ICU admission age, years  (Median, IQR) | 4.2  [1.0, 12.5] | 6.8  [2.0, 13.8] | 3.6  [0.8, 10.5] | 7.4  [2.0, 14.5] |
| Sex, Female, n (%) | 210 (49%) | 224 (41%) | 636 (47%) | 315 (43%) |
| Race, n (%) ^a^ |  |  |  |  |
| White or Caucasian | 226 (53%) | 332 (60%) | 627 (46%) | 421 (57%) |
| Black or African American | 68 (16%) | 87 (16%) | 288 (21%) | 257 (35%) |
| Asian | 5 (1%) | 36 (7%) | 53 (4%) | 2 (<1%) |
| Native Hawaiian or Pacific Islander | 0 (0%) | 5 (1%) | 1 (<1%) | 0 (0%) |
| American Indian or Alaskan Native | 0 (0%) | 4 (1%) | 2 (<1%) | 2 (<1%) |
| Other or Multiple Responses | 31 (7%) | 74 (13%) | 333 (25%) | 15 (2%) |
| Unknown or Refused | 97 (23%) | 15 (3%) | 52 (4%) | 38 (5%) |
| Ethnicity, n (%) ^a^ |  |  |  |  |
| Hispanic or Latino | 31 (7%) | 119 (22%) | 236 (17%) | 83 (11%) |
| Not Hispanic or Latino | 292 (68%) | 263 (48%) | 1046 (77%) | 624 (85%) |
| Other, Unknown, or Refused | 104 (24%) | 171 (31%) | 74 (5%) | 28 (4%) |
| ICU Length of stay, hours  (Median, IQR) | 43.7  [20.6, 106.4] | 45.7  [25.0, 78.9] | 54.8  [29.0, 118.3] | 35.0  [21.3, 69.7] |
| Hospital Disposition ^b^ |  |  |  |  |
| Home or Self Care | 362 (93.3%) | 477 (93.7%) | 1175 (93.0%) | 648 (93.9%) |
| Another Facility or Other | 15 (3.9%) | 28 (5.5%) | 64 (5.1%) | 18 (2.6%) |
| Expired | 11 (2.8%) | 4 (0.7%) | 24 (1.9%) | 24 (3.5%) |

^a^ Race and ethnicity are based on electronic health record extracted values and may not reflect patient self-reporting or be inclusive of all available categories.

^b^ Hospital disposition denominator is based on hospitalizations and not ICU admissions

^a^ Race and ethnicity are based on electronic health record extracted values and may not reflect patient self-reporting or be inclusive of all available categories. ^b^ Hospital disposition denominator is based on hospitalizations and not ICU admission, given the fact that patients could have more than one ICU admission during a single hospitalization. ICU-Intensive care unit. IQR-interquartile range.

**Supplementary Table S2**. Baseline Fluid Metric Comparisons by Site, Including Urine Occurrences

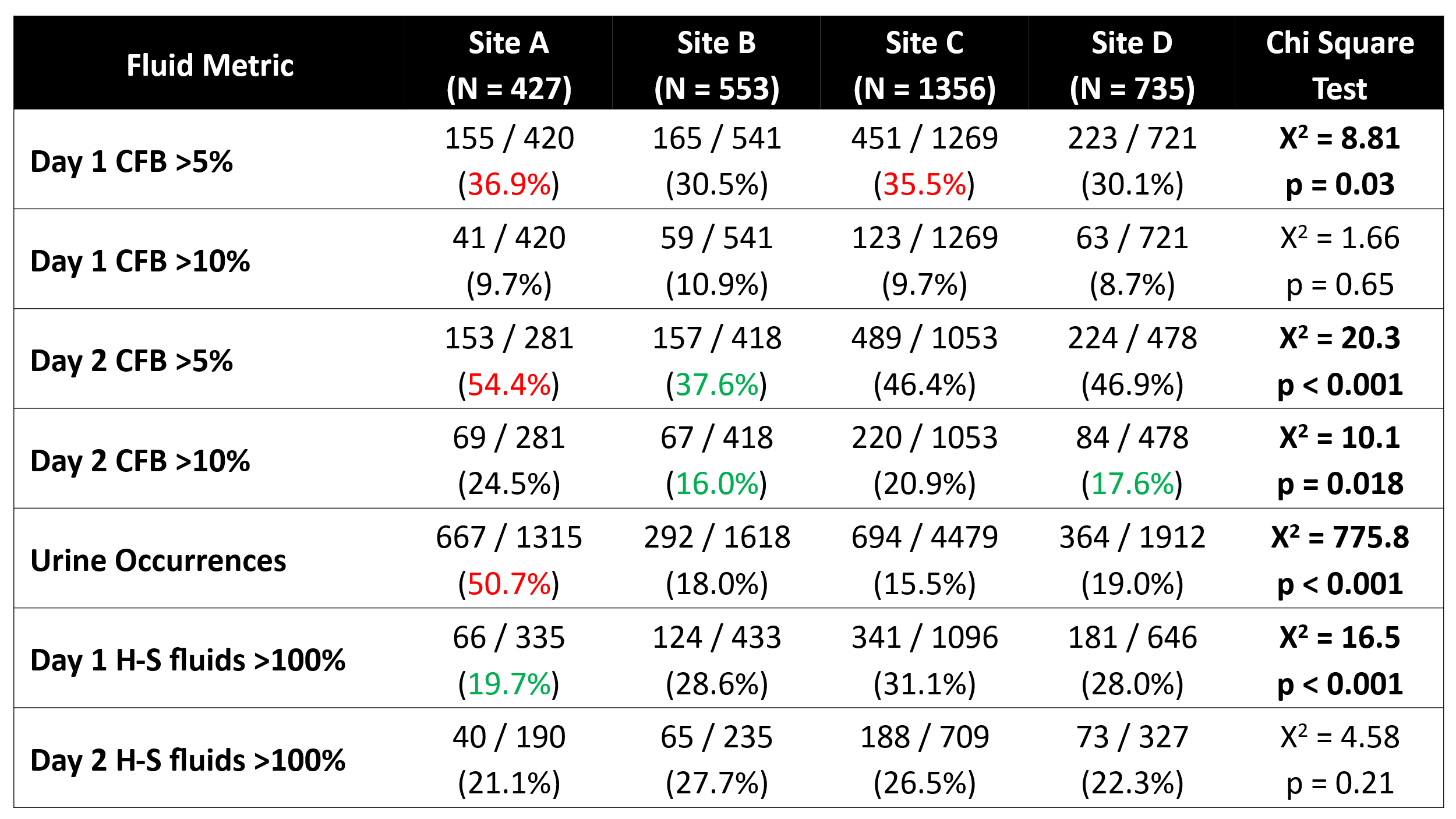

Cumulative fluid balance (CFB), urine occurrence measurements, and IVF intake above H-S calculation for each site. Site-specific sample size in header row reflects total ICU admissions over the study period. Each cell reports # of patient-days (%) meeting fluid metric. Cell-specific denominator reflects total number of ICU patients present on that day of ICU stay, or during any day if no day is specified. CFB measures include patients with and without urine occurrence measurements. H-S measures only include patients with IVF intake. We report row-wise chi-square test of independence p-value to examine relation between fluid metric and site. P values <0.05 are represented by bolding. CFB-cumulative fluid balance, IVF-intravenous fluid. H-S- Holliday-Segar.

**Supplementary Table S2**. Barriers and Opportunities for Accurate Fluid Balance Recording

| **Most Frequent Barriers to Accurate FB Capture** | **Site A**  **N=23** | **Site B**  **N=32** | **Site C**  **N=37** | **Site D**  **N=41** | **Mean Total** |
| --- | --- | --- | --- | --- | --- |
| Patients allowed to PO don’t keep track of oral intake | 14 (61) | 26 (81) | 23 (62) | 31 (76) | 70% |
| Ambulatory patients do not save urine/stool | 11 (48) | 21 (66) | 20 (54) | 25 (61) | 57% |
| Diapers thrown away before weighing | 8 (35) | 14 (44) | 18 (49) | 11 (27) | 39% |
| Unclear process for charting medication flushes | 5 (22) | 8 (25) | 9 (24) | 16 (39) | 28% |
| Too many other tasks de-prioritize FB capture | 8 (35) | 7 (22) | 12 (32) | 11 (27) | 29% |
| Patient acuity precludes strict I/O tracking | 3 (13) | 5 (16) | 7 (19) | 11 (27) | 19% |
| Technical challenges with pump integration | 5 (22) | 2 (6) | 6 (16) | 3 (7) | 13% |
| Enteral intake is less important to track | 3 (13) | 1 (3) | 0 (0) | 2 (5) | 5% |
| Perception that FB is not important/worth the time | 1 (4) | 1 (3) | 4 (11) | 1 (2) | 5% |
| Other | 5 (22) | 3 (9) | 2 (5) | 5 (12) | 12% |
| **Achievable Opportunities to Improve Accurate FB Capture** |  |  |  |  |  |
| Standardized recording of flush volumes | 15 (65) | 23 (72) | 18 (49) | 29 (71) | 64% |
| Standardized flushes, volumes per medication | 10 (43) | 20 (63) | 9 (24) | 22 (54) | 46% |
| Collaboration with other units to add fluids into flowsheets | 9 (39) | 19 (59) | 22 (59) | 19 (46) | 51% |
| Consistent bed zero-ing for daily weights | 12 (52) | 12 (38) | 23 (62) | 9 (22) | 43% |
| Maintaining foley catheters longer | 4 (17) | 9 (28) | 3 (8) | 16 (39) | 23% |
| Larger scales to allow weighing of adult diapers/bed sheets | 8 (35) | 5 (16) | 10 (27) | 12 (29) | 27% |
| Alternatives to foley catheters (e.g., pure wick, condom catheters) | 5 (22) | 4 (13) | 17 (46) | 8 (20) | 25% |
| Other | 1 (4) | 1 (3) | 1 (3) | 3 (7) | 4% |
| **Most Feasible Method for Accurate FB Capture** |  |  |  |  |  |
| Setting a total fluid goal on rounds, discussing ways to meet it | 15 (65) | 17 (53) | 19 (51) | 19 (46) | 54% |
| Strict I/O without occurrence measurements | 6 (26) | 12 (38) | 13 (35) | 12 (29) | 32% |
| Daily weights | 2 (9) | 2 (6) | 5 (14) | 10 (24) | 13% |

Results of surveys completed by bedside nurses. First 2 questions asked for the respondent’s top 3 choices. The last question asked for the single best answer. FB-fluid balance, PO-oral, I/O-intake/output
